# A proteomic survival predictor for COVID-19 patients in intensive care

**DOI:** 10.1101/2021.06.24.21259374

**Authors:** Vadim Demichev, Pinkus Tober-Lau, Tatiana Nazarenko, Simran Kaur Aulakh, Harry Whitwell, Oliver Lemke, Annika Röhl, Anja Freiwald, Mirja Mittermaier, Lukasz Szyrwiel, Daniela Ludwig, Clara Correia-Melo, Lena J. Lippert, Elisa T. Helbig, Paula Stubbemann, Nadine Olk, Charlotte Thibeault, Nana-Maria Grüning, Oleg Blyuss, Spyros Vernardis, Matthew White, Christoph B. Messner, Michael Joannidis, Thomas Sonnweber, Sebastian J. Klein, Alex Pizzini, Yvonne Wohlfarter, Sabina Sahanic, Richard Hilbe, Benedikt Schaefer, Sonja Wagner, Felix Machleidt, Carmen Garcia, Christoph Ruwwe-Glösenkamp, Tilman Lingscheid, Laure Bosquillon de Jarcy, Miriam S. Stegemann, Moritz Pfeiffer, Linda Jürgens, Sophy Denker, Daniel Zickler, Claudia Spies, Andreas Edel, Nils B. Müller, Philipp Enghard, Aleksej Zelezniak, Rosa Bellmann-Weiler, Günter Weiss, Archie Campbell, Caroline Hayward, David J. Porteous, Riccardo E. Marioni, Alexander Uhrig, Heinz Zoller, Judith Löffler-Ragg, Markus A. Keller, Ivan Tancevski, John F. Timms, Alexey Zaikin, Stefan Hippenstiel, Michael Ramharter, Holger Müller-Redetzky, Martin Witzenrath, Norbert Suttorp, Kathryn Lilley, Michael Mülleder, Leif Erik Sander, PA-COVID-19 Study group, Florian Kurth, Markus Ralser

**Author notes:** To whom Correspondence should be addressed: Florian Kurth M.D. These authors contributed equally (Vadim Demichev, Pinkus Tober-Lau). **PA-COVID-19 Study group, Charité – Universitätsmedizin Berlin** Malte Kleinschmidt, Katrin M. Heim, Belén Millet, Lil Meyer-Arndt, Nils B. Müller, Ralf H. Hübner, Tim Andermann, Jan M. Doehn, Bastian Opitz, Birgit Sawitzki, Daniel Grund, Peter Radünzel, Mariana Schürmann, Thomas Zoller, Fridolin Steinbeis, Florian Alius, Philipp Knape, Astrid Breitbart, Yaosi Li, Felix Bremer, Panagiotis Pergantis, Susanne Fieberg, Anne Wetzel, Moritz Müller-Plathe, Timur Özkan, Carola Misgeld, Dirk Schürmann, Bettina Temmesfeld-Wollbrück, Britta Stier, Martin Möckel, Jan A. Graaw, Victor Wegener, Marc Kastrup, Felix Balzer, Daniel Wendisch, Sophia Brumhard, Sascha S. Haenel, Philipp Georg, Claudia Conrad, Kai-Uwe Eckardt, Lukas Lehner, Jan M. Kruse, Carolin Ferse, Roland Körner, Andreas Edel, Steffen Weber-Carstens, Alexander Krannich, Saskia Zvorc, Linna Li, Uwe Behrens, Sein Schmidt, Maria Rönnefarth, Christina Pley, Claudia Fink, Chantip Dang-Heine, Robert Röhle, Emma Lieker, Christian Wollboldt, Yinan Wu, Georg Schwanitz, Constanze Lüttke, Denise Treue, Michael Hummel, Victor M. Corman, Christian Drosten, Christof von Kalle.

## Abstract

Global healthcare systems are challenged by the COVID-19 pandemic. There is a need to optimize allocation of treatment and resources in intensive care, as clinically established risk assessments such as SOFA and APACHE II scores show only limited performance for predicting the survival of severely ill COVID-19 patients. Comprehensively capturing the host physiology, we speculated that proteomics in combination with new data-driven analysis strategies could produce a new generation of prognostic discriminators. We studied two independent cohorts of patients with severe COVID-19 who required intensive care and invasive mechanical ventilation. SOFA score, Charlson comorbidity index and APACHE II score were poor predictors of survival. Plasma proteomics instead identified 14 proteins that showed concentration trajectories different between survivors and non-survivors. A proteomic predictor trained on single samples obtained at the first time point at maximum treatment level (i.e. WHO grade 7) and weeks before the outcome, achieved accurate classification of survivors in an exploratory (AUROC 0.81) as well as in the independent validation cohort (AUROC of 1.0). The majority of proteins with high relevance in the prediction model belong to the coagulation system and complement cascade. Our study demonstrates that predictors derived from plasma protein levels have the potential to substantially outperform current prognostic markers in intensive care.

**Trial registration:** German Clinical Trials Register DRKS00021688

## Introduction

The COVID-19 pandemic has brought health systems around the globe to the brink of collapse. Capacities for intensive care treatment of patients with organ failure have reached their limits in many regions with intense SARS-CoV-2 transmission and were often central to political decisions regarding restrictions on public life, e.g. through contact restrictions or lock-downs. Various models for classification of disease severity and for prediction of clinical trajectories and outcome have been developed for COVID-19, based on laboratory measurements, clinical scores, imaging, and omics technologies [1–4]. These pointed to the importance of specific immune cells, inflammatory and antiviral cytokines and chemokines, as well as the coagulation cascade in COVID-19 disease progression [4–12]. They predict the risk of the future need for mechanical ventilation in the heterogeneous group of patients at early time points, e.g. at admission to the hospital, when clinical parameters and biomarkers differ substantially between mildly affected and severely ill patients [1–4,13].

Treatment decisions within the most severely ill patients, for instance whether a patient should be treated with extracorporeal membrane oxygenation (ECMO), have a major impact on resources. Currently such decisions are often based primarily on patient’s age, comorbidities, and established intensive care prognosis models, such as the Sequential Organ Failure Assessment (SOFA) or Acute Physiology and Chronic Health Evaluation (APACHE II), which assess the patient on the basis of a combination of established clinical and laboratory risk parameters [14,15]. The predictive values of both SOFA and APACHE II for the most critical forms of COVID-19 are limited [16–18], creating a diagnostic gap and imminent need for reliable predictors, specifically validated in severely ill COVID-19 patients, to guide and tailor efforts in treating these critically ill patients. Indeed, classifying clinical trajectories within more homogeneous groups such as WHO grade 7 patients is considerably more difficult to achieve than a molecular severity classification that distinguishes mild from severe patients; physiological and molecular differences are less pronounced within the same than between different severity groups. As a consequence, the relative impact of confounders and random environmental factors on molecular and physiological parameters for clinical decision making is stronger.

Plasma proteomics holds the promise of integrating the genetic background of an individual with their life history, physiological, nutritional, and demographic parameters, and hence, have the potential to form the foundation of a new generation of predictors [19–23]. Among the spectrum of proteomic technologies available, mass spectrometry has the appeal that once markers are identified, they allow for the direct generation of targeted panel assays measurable by selective reaction monitoring (SRM), simplifying their implementation into clinical routine. Recently, new mass spectrometry based proteomic technologies have been developed to increase throughput and measurement precision, so that the path from discovery to application is simplified [5,24–27].

We studied proteomes of two well characterized cohorts of the most severely ill patients with COVID-19 in two independent health care centers (Charité-Universitätsmedizin, Berlin Germany, and Medical University of Innsbruck, Austria) who gave informed consent to deep clinical and molecular phenotyping [13,18,28], We found 14 protein concentration trajectories that distinguish survivors from non-survivors. Moreover, a machine learning (ML) model, based on parenclitic networks, generated accurate prognosis on single time point samples that were collected once the patient reached the maximum treatment level, in median, 39 days before outcome. The ML predictor substantially outperformed established clinical risk scores.

## Results

The exploratory cohort used for marker identification and model generation consisted of the 50 most severely ill COVID-19 patients out of a cohort of 168 patients with varying disease severity, treated between 15 March and 16 September 2020 at Charité University Hospital, Berlin, Germany (**Fig. 1A**) [13,18,28]. All 50 patients required intensive care with invasive mechanical ventilation plus additional organ support such as renal replacement therapy (RRT), ECMO, or vasopressors, corresponding to grade 7 on the WHO Ordinal Scale for Clinical Improvement. Patients with limitations of therapy according to their wish were excluded. There were no treatment restrictions due to shortages of intensive care capacity at the time of this patient cohort. Of the 50 patients, 36 (72%) required RRT, 19 (38%) patients were treated with ECMO, and 16 (32%) patients were treated with both RRT and ECMO. Fifteen (30%) patients died. Median time of hospitalization in survivors was 63 days (n=35, IQR 44-89). Median time from admission to death was 28 days (n=15, IQR 16-43). Patient characteristics are shown in Supplementary table 1.

**Figure 1.**
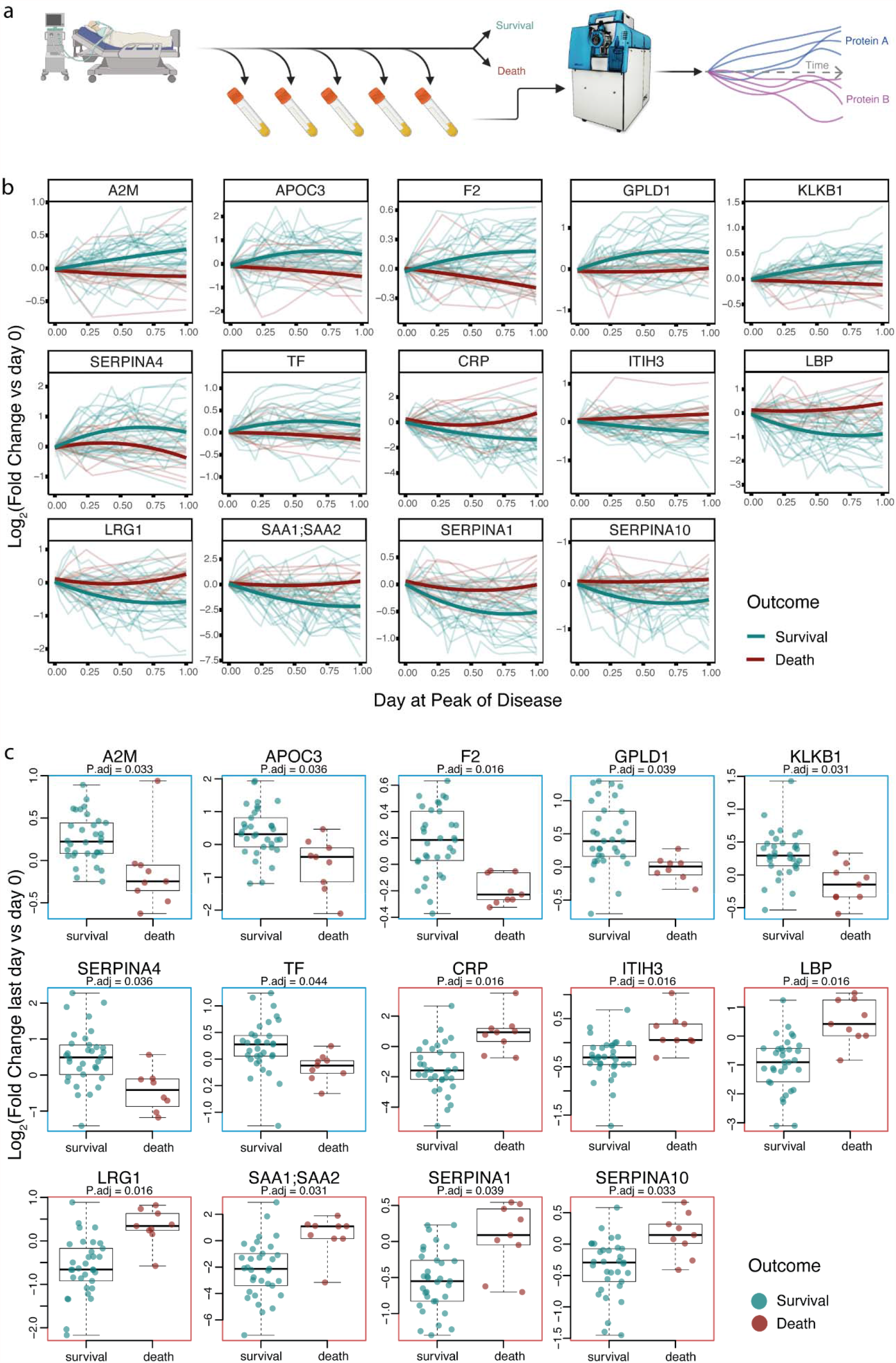
Protein concentration trajectories that differentiate survivors of critical COVID-19 from non-survivors. **a)** Patients with PCR-confirmed COVID-19 treated at Charité University Hospital Berlin, Germany, were sampled longitudinally, to generate high-resolution time series for 321 protein quantities. In parallel, precise clinical phenotyping was performed, including recording of intensive care and disease severity scores, treatment parameters, and outcome (PA-COVID 19 Data ressource, [13]). **b)** Protein level trajectories over time (FDR < 0.05), for which time-dependent concentration changes (y-axis: log2 fold change) during the peak of the disease differentiate survivors from non-survivors in critically ill patients (Methods). **c)** as **b)** but expressed as a boxplots (log2 fold change last vs first day).

The Charlson Comorbidity Index [29,30], performed poorly in classifying survivors from non-survivors by AUROC values of 0.63 (P = 0.16, **Fig. 2A**). From a time-resolved data resource for the PA-COVID-19 study, spanning over a compendium of clinical parameters, plasma proteomes, cell counts, enzyme activities, and outcomes [13], we further determined the SOFA and APACHE II scores. These scores, too, could not confidently distinguish survivors from non-survivors (**Fig. 2A**, AUROC = 0.68, P = 0.05 for APACHE II score at ICU admission and AUROC = 0.65, P = 0.11 for SOFA score at the time of first sampling at WHO grade 7).

Studying the plasma proteomes [13] we found 78 proteins the concentration of which significantly changed during the patients’ disease course. Out of these proteins, 14 were found to change differently over time for survivors and non-survivors (**Fig. 1B, C)**. This included a significant increase in inflammatory proteins over time (SAA1, SAA2, CRP, ITIH3, LRG1, SERPINA1, SERPINA10 and LBP) in patients with fatal outcomes, and a corresponding decrease in survivors. Likewise, a decrease in anti-inflammatory proteins (SERPINA4, A2M) was noted in non-survivors but not in survivors, indicating a persistent pro-inflammatory signature in the former. Similarly, two key molecules in the coagulation system, thrombin (F2) and plasma kallikrein (KLKB1), known to be decreased in severe COVID-19 [11,13], further decreased over time in non-survivors while increasing in survivors.

**Figure 2.**
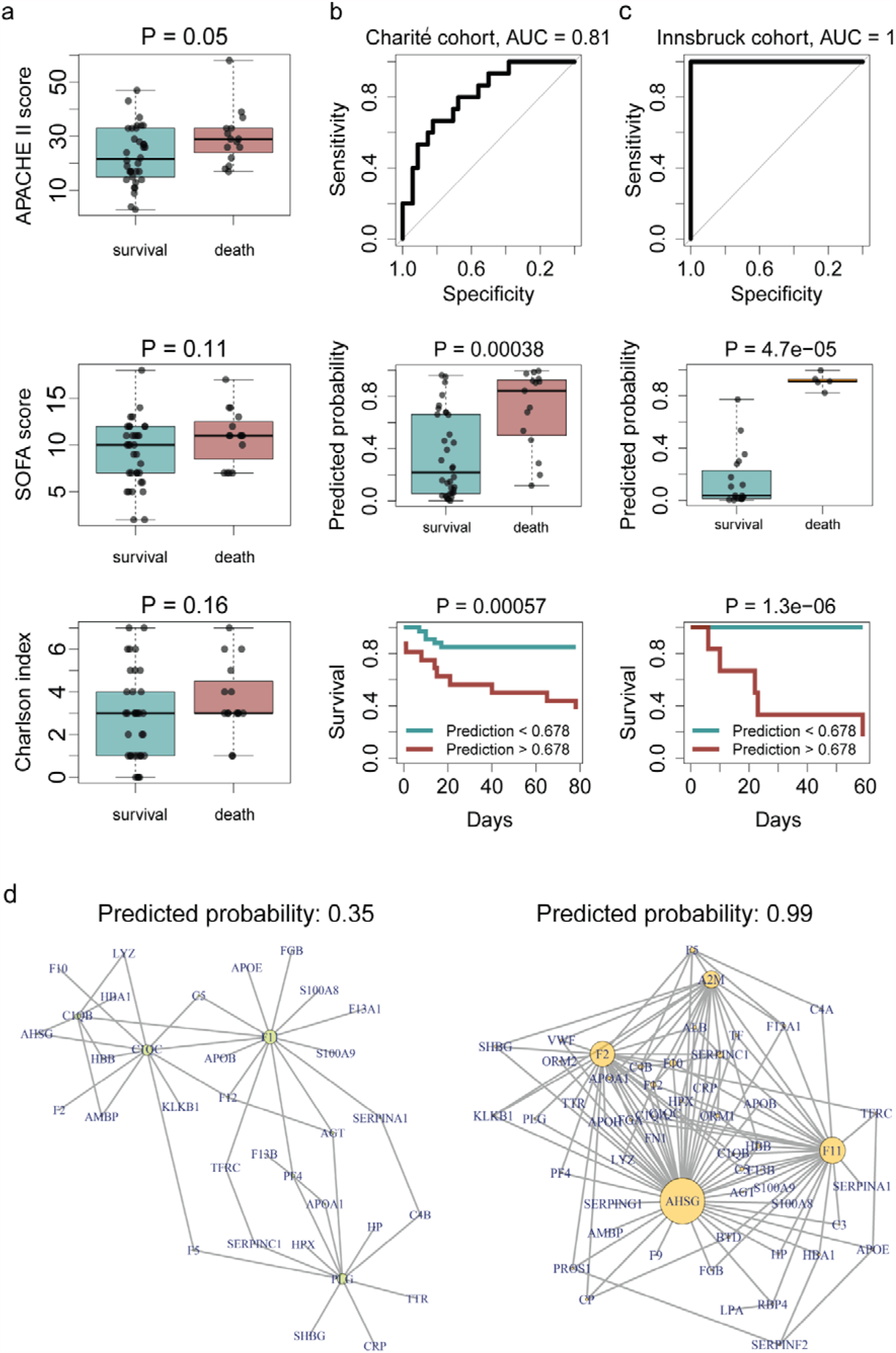
Prediction of survival or death in critically ill patients, from the first sampling time point at intensive care treatment level (WHO grade 7). **a)** Performance of established ICU risk assessment indices (APACHE II, SOFA and Charlson comorbidity index) calculated at the time of ICU admission (APACHE II, Charlson comorbidity index) or at the first time point at WHO grade 7 (SOFA score) in predicting the outcome in critically ill patients. **b)** Prediction of survival or death in critically ill patients using proteomics. A machine learning model based on parenclitic networks (Methods) was trained on the samples of the Charité cohort closest to the time point of treatment escalation during intensive care (start of ECMO, RRT or vasopressors, i.e. WHO grade 7). The performance was assessed on the test samples, which were held out during training. **Upper panel:** The ROC curve indicates correct classification of survival vs non-survival with an AUROC of 0.81. **Middle panel**: The proteomic classifier was used to predict the probability of survival and non-survival, which is significantly different between the groups. **Lower panel**: Kaplan-Meier survival curves using a threshold of predicted probability (0.678) chosen to maximize Youden’s J index (J = sensitivity + specificity - 1). Log-rank test was used to compare survival rates between patients with predicted death risk < 0.678 (black) and > 0.678 (orange). **c) (upper, middle, and lower panels):** The model trained on the Charité cohort, was tested on an independent cohort (Innsbruck). **d)** Exemplary parenclitic networks from two patients in the independent Innsbruck cohort. Edges with weights > 0.5 are shown. Left panel: a network predicting low probability of death in a surviving patient. Right panel: a network predicting high probability of death in a non-survivor.

For diagnostic purposes and treatment decisions time series data is however difficult to implement. We therefore explored the potential of using the earliest sample obtained at the maximum treatment level (WHO grade 7), i.e. at a time point critical for decisions about escalation of treatment, to predict the clinical outcomes (survival). The median time until the outcome was 39 (IQR 16 - 64) days. We established a machine learning model based on parenclitic networks, a graph-based approach in which networks representing the deviation of an individual from the population are derived [31,32]. The networks are generated (Methods) by considering every pair of analytes (proteins) individually and calculating the respective edge weight as the estimated probability of fatal outcome based on this pair of proteins. Predictive models are then generated by considering the topological differences between networks from individual cases (non-survivors vs. survivors). We achieved high prediction accuracy on the test subjects, who were excluded when training the machine learning model (in a cross-validation fashion, see Methods), with AUC = 0.81 for the receiver-operating characteristic (ROC) curve (**Fig. 2B**). Out of the 25 proteins with the highest relevance in the parenclitic model, 15 are components of the coagulation system and 8 proteins belong to the complement cascade (Supp. Table 2).

To independently validate the proteomic predictor, we examined its performance on an independent cohort of 24 patients with critical COVID-19 from Austria (survival n=19, death n=5, median time between sampling and outcome 22 days, interquartile range 15 - 42 days) (‘Innsbruck’ cohort, Methods). Despite the validation cohort originating from a different hospital and health care system, the machine learning model demonstrated high predictive power on this independent cohort (AUROC = 1.0, P = 0.00038, **Fig. 2C**). Using the cutoff value for survival prediction derived from the Charité cohort, the model correctly predicted the outcome for 18 out of 19 patients who survived and for 5 out of 5 patients who died in this independent ‘Innsbruck’ cohort.

## Discussion

The prognostic value of several biomarkers (e.g. CRP, IL-6, ferritin) and clinical scores for predicting disease progression in COVID-19 at early disease stages, e.g. at hospital admission, is now well established [33,34]. For the comparatively homogeneous subgroup of severely ill patients already requiring mechanical ventilation and additional organ support, prediction of future disease trajectories and outcome (survival or death) is by far more challenging and only limited data exist [16,35,36]. Moreover, clinical severity scores are often not validated for unconscious patients and laboratory measurements are frequently confounded by intensive care treatment. Outcome of ICU patients may further be critically determined by resource constraints, the varying level of experience with organ replacement therapies or the rates of superinfection, rendering prediction complex [36]. On the other hand, patients in intensive care units, and particularly those in need of special organ replacement therapies such as ECMO, require a disproportionately large share of resources compared to other patients, so decisions to initiate such therapies should be based on the best information and assessment possible. Prognostic tools in critically ill patients are hence of crucial importance to guide and tailor the treatment efforts. This is particularly true in a situation when health care systems are overstrained.

Previously, we and others investigated plasma proteome alterations in COVID-19 [5–7,9,11,13], which show a remarkable ability to classify the severity of disease. New proteomic platform technologies have significantly gained precision and throughput compared to their predecessors, rendering the application of multivariate regression models more effective and bringing them increasingly close to routine clinical use [5]. Importantly, even without platform technologies, biomarkers identified in proteomic profiles can be translated into clinical use, e.g. by using standard techniques such as selective reaction monitoring (SRM) or ELISA.

Here, we show that an increase in specific inflammatory and acute phase proteins over time (e.g., SAA1;SAA2, CRP, ITIH3, LRG1, SERPINA1, and LBP) is associated with the risk of death from COVID-19, while an increase of kallikrein (KLKB1), kallistatin (SERPINA4), thrombin (F2), apolipoprotein C3 (APOC3), GPLD1, and the protease inhibitor A2M, is associated with survival. Kallikrein is involved in the blood coagulation system, fibrinolysis, and the complement cascade, three systems known to be dysregulated in COVID-19 [37–39]. It mediates the cleavage of kininogen to bradykinin and des-Arg^9^-bradykinin, a potent vasoactive peptide which is counter-regulated by ACE2, the cell entry receptor for SARS-CoV-2. Since the loss of ACE2 in COVID-19 supposedly leads to an imbalance of bradykinins, inhibition of the kallikrein-kinin system has been discussed as treatment strategy in COVID-19 [40–42]. This hypothesis is not supported by our data, which indicate improved prognosis with increasing kallikrein levels. Kallikrein is counterbalanced by kallistatin, which equally increased over time in survivors in our study population, thereby potentially equilibrating the increase in the kinin-kallikrein system. Kallistatin is known for pleiotropic effects in vascular repair, endothelial function, and inflammation [43] and possesses protective properties in acute lung injury. According to our data kallistatin should be considered as a potential candidate for clinical testing in critical COVID-19 [44].

While prognostic assessments based on repeated measurements over time allow for treatment monitoring, including evaluation of experimental therapies in clinical trials, prognostic measurements from single time points are particularly valuable for timely patient management and resource allocation. We therefore employed a machine learning model to integrate proteomic measurements from the first time point at WHO grade 7, i.e. invasive mechanical ventilation and additional organ support therapy, in order to derive prognosis of outcome. We achieve high prognostic values, both in the exploratory cohort, as well as in a fully independent cohort.

The majority of proteins with the highest relevance for the machine learning predictor were components of the coagulation system and the complement cascade (Supp. Table 2). Both systems are known to be crucial for treatment and disease courses for severely ill COVID-19 patients [8,9]. This is particularly well illustrated by recent data from a multi-platform clinical trial indicating that a substantial proportion of patients with severe COVID-19 develop thromboembolic events despite therapeutic anticoagulation [45,46]. The protein with the highest relevance in our model is Fetuin-A (AHSG), which is known to be strongly downregulated in severe COVID-19 [9,13]. Of note, genetic polymorphisms associated with higher AHSG plasma concentrations were found to be protective in SARS-CoV-1 infection [47]. One important function of AHSG is regulation of inflammation through deactivation of macrophages [48], and there is emerging evidence that macrophages play a key role in pulmonary inflammation and dysfunction in COVID-19 [10,49–51].

In summary, we have leveraged the power of the proteome to address a problematic diagnostic gap in the prognosis of the most critical form of COVID-19, that is not covered by established clinical assessments, such as the SOFA or APACHE II score. We show that the proteome accurately predicts survival in critically ill patients with COVID-19, from samples that were collected 39 days in median before the outcome. The majority of proteins with high relevance in the model are components of the coagulation system and complement cascade, highlighting their critical role in progression and outcome of most severe COVID-19.

## Methods

### Charité patient cohort and clinical data

Patients included in this analysis are a sub-cohort of the Pa-COVID-19 study conducted at Charité - Universitätsmedizin Berlin, a prospective observational cohort study on the pathophysiology of COVID-19 as described previously [18,28]. All patients with PCR-confirmed SARS-CoV-2 infection that progressed to critical disease (WHO grade 7, i.e. invasive mechanical ventilation and additional organ support), were eligible for inclusion. Exclusion criteria included refusal to provide informed consent by the patient or a legal representative, and any condition prohibiting serial biosampling. Patients were treated according to current clinical guidelines. Patients for whom limitation of therapy was decided according to the patient’s wish were excluded from analysis. In three further cases, limitation of therapy was decided at a later time point according to the patient’s presumed wish and predictably unfavorable outcome. All other patients received maximum intensive care treatment including organ replacement therapies at the discretion of the responsible physicians. One patient (ID 135), who was still hospitalized and clinically improving 5 months after admission, was classified as a survivor. Patients still in critical condition 5 months after admission were excluded due to uncertain outcome.

Biosampling of EDTA plasma for proteome measurement was performed up to 3 times per week after inclusion. Disease severity was assessed according to the WHO ordinal scale for clinical improvement (World Health Organisation 2020). Clinical data were in SecuTrial®. Pseudonymized data exported from SecuTrial® were processed using JMP Pro 15 (SAS Institute Inc., Cary, NC, USA).

### Innsbruck Patient cohort and clinical data

Serum samples from patients admitted to the intensive care unit at the Department of Medicine, University Hospital of Innsbruck with PCR-confirmed severe COVID-19 were collected within the first days (median 7.5, IQR 5-12) after admission, and written informed consent was obtained. Patients were treated according to national guidelines. The study was approved by the local ethics research committee EK-Nr. 1107/2020, and EK-Nr. 1103/2020 for follow-up.

### Statistical analysis and multiple-testing correction

Statistical testing on proteomic and diagnostic data [13] was performed in the R environment for statistical computing, version 3.6.0 [52]. All protein measurements were first log2-transformed and only protein groups matched to at least three different peptides were considered. Quantities of gene products corresponding to open reading frames IGxx (i.e. different types of immunoglobulin chains) were summed together to generate quantities representative of the overall levels of immunoglobulin classes (IGHVs, IGLVs, etc). Significance testing for equal medians was performed using the Mann-Whitney U test, as implemented in the “wilcox.test” function of the “stats” R package. Multiple-testing correction was performed using the Benjamini-Hochberg false discovery rate controlling procedure [53], implemented in the “p.adjust” function of the “stats” R package. Adjusted p-values below 0.05 were considered significant.

### Identifying omics trajectories that are predictive of survival at the peak period of the disease

For each omics feature, the difference between its log2-levels at the last and the first sampling timepoints during the peak period of the disease was considered. This period was defined as the time when the patient was receiving the most intensive treatment during their stay in hospital, that is the time when the patient was at WHO grade 6 or 7. The distribution of this difference between survivors and non-survivors was compared using the Mann-Whitney U test. Only non-DNI patients with known outcome were included.

### Prediction of survival

The first time point measured at the WHO grade 7 was selected per patient. To reduce the feature space used as input for the machine learning model, we limited it to the quantities of 57 proteins which are FDA-approved biomarkers with MRM assays available [54] and which were quantified with at least three different peptides in this study. Missing values were imputed using minimal value imputation, and the data were standardized.

Machine learning was carried out using the parenclitic networks approach [31,55]. Briefly, during training, for each pair of features, a radial SVM classifier is trained (using the svm() function from the “e1071” R package with default settings). For each sample, a network is then built, wherein vertices correspond to features and the edge weight is the death probability as predicted by the SVM classifier. Maximum, mean and standard deviation of the edge weights, as well as the numbers of edges with weights greater than 0.5 (i.e. fatal outcome is predicted) and nodes with at least one such edge are calculated. A LASSO classification model (alpha = 0.01) is then constructed on these 5 features using the glmnet() function of the “glmnet” [56] R package with default settings.

For the assessment of the classifier performance (Charité cohort), a cross-validation method was applied in the following way: the prediction was made for each sample by excluding (withholding) it from the dataset along with two other samples (chosen randomly with the constraint that out of 3 samples one corresponds to a non-survivor and two to survivors), training the classifier on the remaining (independent) samples and then generating predictions for the withheld samples using the trained model. Such a leave-3-out partition was generated randomly 50 times and the predictions for each sample were averaged. For the assessment of the performance on an independent dataset (Innsbruck cohort), the classifier was trained on all the Charité samples and used to estimate the probabilities of fatal outcome on the Innsbruck cohort. The source code is provided in supplementary materials.

The ‘relevance’ scores for proteins in the parenclitic model were calculated as Kleinberg’s authority centrality scores for the respective vertices in the “generalizing network”. This network was generated by (i) replacing edge weights greater than 0.5 with 1.0 and weights less than 0.5 with 0.0 in the networks corresponding to non-survivors and (ii) averaging the resulting networks.

## Supporting information

Supplementary data and machine learning code

## Data Availability

The protein quantities table along with the associated metadata are provided in supplementary materials. All scripts used to train and assess the machine learning models are likewise provided.

## Study approval

The study was approved by the ethics committee of Charité - Universitätsmedizin Berlin (EA2/066/20) and conducted in accordance with the Declaration of Helsinki and guidelines of Good Clinical Practice (ICH 1996). Written informed consent was obtained from all patients or legal representatives according to regulations set by the ethics committee of Charité - Universitätsmedizin Berlin. The study is registered in the German and the WHO international registry for clinical studies (DRKS00021688).

## Acknowledgements

We thank Jan-David Manntz (Beckman, Germany) for help with the Biomek i7, Robert Lane, Jean-Baptiste Vincedent and Nick Morrice (SCIEX) for help with the TripleTOF 6600. This work was supported by the Berlin University Alliance (501_Massenspektrometrie, 501_Linklab, 112_PreEP_Corona_Ralser), by UKRI/NIHR through the UK Coronavirus Immunology Consortium (UK-CIC), the BMBF/DLR Projektträger (01KI20160A, 01ZX1604B, 01KI20337, 01KX2021), Charité-BIH Centrum für Therapieforschung (BIH_PA_covid-19_Ralser), the BBSRC (BB/N015215/1, BB/N015282/1), the Francis Crick Institute, which receives its core funding from Cancer Research UK (FC001134), the UK Medical Research Council (FC001134), and the Wellcome Trust (FC001134 and IA 200829/Z/16/Z). The work was further supported by the Ministry of Education and Research (BMBF), as part of the National Research Node ‘Mass spectrometry in Systems Medicine (MSCoresys), under grant agreement 031L0220A. Leif Erik Sander is supported by the German Research Foundation (DFG, SFB-TR84 114933180) and by the Berlin Institute of Health (BIH), which receives funding from the Ministry of Education and Research (BMBF). Martin Witzenrath is supported by grants from the German Research Foundation, SFB-TR84 C06 and C09, by the German Ministry of Education and Research (BMBF) in the framework of the CAPSyS (01ZX1304B), CAPSyS-COVID (01ZX1604B), SYMPATH (01ZX1906A) and PROVID project (01KI20160A) and by the Berlin Institute of Health (CM-COVID). Stefan Hippenstiel is supported by the German Research Foundation (DFG, SFB-TR84 A04 and B06), and the BMBF (PROVID, and project 01KI2082). Norbert Suttorp is supported by grants from the German Research Foundation, SFB-TR84 C09 und Z02, by the German Ministry of Education and Research (BMBF) in the framework of the PROGRESS 01KI07114. The study was further supported by Wellcome Trust (200829/Z/16/Z). The Generation Scotland study received core support from the Chief Scientist Office of the Scottish Government Health Directorates (CZD/16/6) and the Scottish Funding Council (HR03006), and is now supported by the Welcome Trust (216767/Z/19/Z). Archie Campbell is funded by HDR UK and the Wellcome Trust (216767/Z/19/Z). Caroline Hayward is supported by an MRC University Unit Programme Grant (MC_UU_00007/10) (QTL in Health and Disease). Riccardo Marioni is supported by an Alzheimer’s Research UK project grant (ARUK-PG2017B-10). H. Whitwell, JF. Timms, A. Zaikin and T. Nazarenko are supported by a Medical Research Council grant (MR/R02524X/1) and H. Whitwell, A. Zaikin and O. Blyuss by the Ministry of Science and Higher Education agreement No. 075-15-2020-808. H. Whitwell is supported by the National Institute for Health Research (NIHR) Imperial Biomedical Research Centre (BRC). J. Timms is supported by the National Institute for Health Research (NIHR) UCLH/UCL Biomedical Research Centre. Mirja Mittermaier is a participant in the BIHCharité Digital Clinician Scientist Program funded by the Charité – Universitätsmedizin Berlin, the Berlin Institute of Health, and the German Research Foundation (DFG). Markus A. Keller is supported by the Austrian Science Funds (FWF; P33333) and the Austrian Research Promotion Agency (FFG, #878654). Figures were created with biorender.com.

---

**Supplementary Table 1.** Baseline, treatment, and outcome characteristics of patient cohort with severe COVID-19 receiving maximum therapy at Charité - University hospital Berlin.

|  | all patients |  | survived |  | deceased |  |
| --- | --- | --- | --- | --- | --- | --- |
| **Number of patients** | 50 | 100 % | 35 | 70 % | 15 | 30 % |
| **Sex** |  |  |  |  |  |  |
| Female | 15 | 30 % | 12 | 34 % | 3 | 20 % |
| Male | 35 | 70 % | 23 | 66 % | 12 | 80 % |
| **Age, years (Median, IQR)** | 62 | [54 - 73] | 61 | [54 - 69] | 69 | [56 - 76] |
| ≥ 65 | 22 | 44 % | 12 | 34 % | 10 | 67 % |
| **BMI (kg/m<sup>2</sup>, Median, IQR), n = 49** | 29.4 | [27.4 - 34.7] | 31.0 | [27.4 - 35.1] | 29.0 | [25.7 - 31.2] |
| < 25 kg/m<sup>2</sup> | 9 | 18 % | 6 | 18 % | 3 | 20 % |
| ≥ 25 kg/m<sup>2</sup> | 40 | 82 % | 28 | 82 % | 12 | 80 % |
| **Pre-existing conditions** |  |  |  |  |  |  |
| Charlson's Comorbidity Index (Median, IQR) | 3 | [1 - 4] | 3 | [1 - 4] | 3 | [3 - 5] |
| CCI <3 | 18 | 36 % | 16 | 46 % | 2 | 13 % |
| CCI ≥3 | 32 | 64 % | 19 | 54 % | 13 | 87 % |
| h/o smoking | 8 | 16 % | 6 | 17 % | 2 | 13 % |
| current smoker | 1 | 2 % | 1 | 3 % | 0 | 0 % |
| **Outpatient medications (Median, IQR)** | 2 | [1 - 4] | 2 | [1 - 4] | 2 | [1 - 3] |
| **Duration of hospital course (days, median, IQR)** | 50 | [33 - 79] | 63 | [44 - 89] | 28 | [16 - 43] |
| Proning | 42 | 84 % | 30 | 86 % | 12 | 80 % |
| RRT | 36 | 72 % | 23 | 66 % | 13 | 87 % |
| ECMO | 19 | 38 % | 12 | 34 % | 7 | 47 % |
| ARDS | 49 | 98 % | 34 | 97 % | 15 | 100 % |
| Sepsis | 29 | 58 % | 18 | 51 % | 11 | 73 % |
| Thromboembolic event | 23 | 46 % | 17 | 49 % | 6 | 40 % |
| Cardiopulmonary resuscitation | 5 | 10 % | 4 | 12 % | 1 | 7 % |
| **Outcome** |  |  |  |  |  |  |
| Deceased (incl. secondary DNR) | 15 | 30 % | - | - | 15 | 100 % |
| Secondary DNR | 3 | 6 % | - | - | 3 | 6 % |
| Requiring new oxygen therapy after discharge* | 10 | 29 % | 10 | 29 % | - | - |
| Requiring new RRT after discharge* | 5 | 14 % | 5 | 14 % | - | - |
| Data are shown in n (%) unless otherwise indicated. IQR - interquartile range, BMI - body mass index, RRT - renal replacement therapy, ECMO - extracorporeal membrane oxygenation, secondary DNR - secondary limitation of therapy in situation of probable unfavorable outcome and according to the presumed patient's wish |  |  |  |  |  |  |
| * Deceased patients not included |  |  |  |  |  |  |

**Supplementary Table 2.**
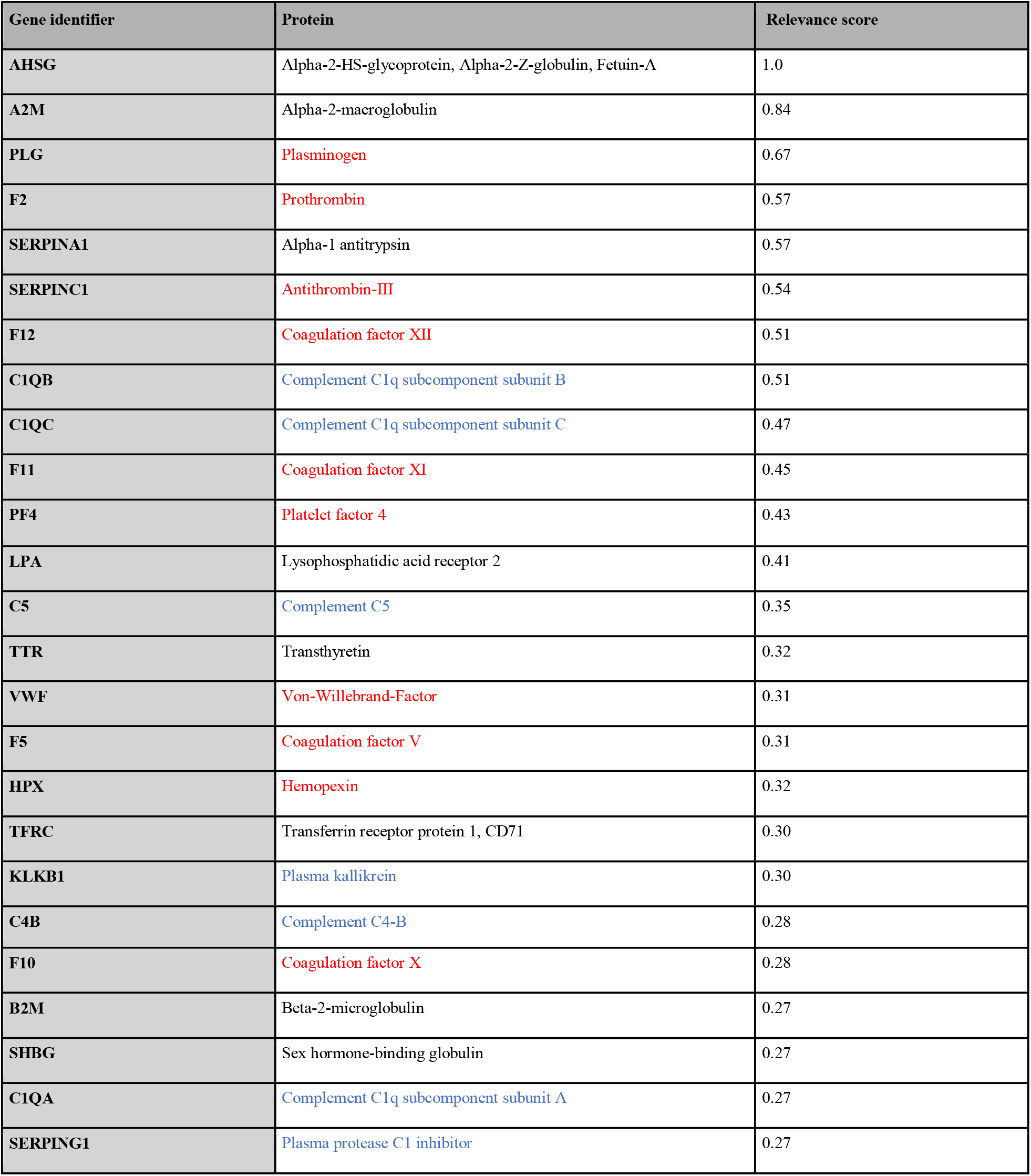
Top 25 proteins included in the machine learning model, ordered by their estimated ‘relevance’ scores (Methods). Red writing indicates proteins involved in the complement system. Blue writing indicates proteins involved in coagulation.

## References

1. Clift AK, Coupland CAC, Keogh RH, Diaz-Ordaz K, Williamson E, Harrison EM, et al. Living risk prediction algorithm (QCOVID) for risk of hospital admission and mortality from coronavirus 19 in adults: national derivation and validation cohort study. BMJ. 2020;371: m3731.

2. Knight SR, Ho A, Pius R, Buchan I, Carson G, Drake TM, et al. Risk stratification of patients admitted to hospital with covid-19 using the ISARIC WHO Clinical Characterisation Protocol: development and validation of the 4C Mortality Score. BMJ. 2020;370. doi:10.1136/bmj.m3339

3. Chassagnon G, Vakalopoulou M, Battistella E, Christodoulidis S, Hoang-Thi T-N, Dangeard S, et al. AI-driven quantification, staging and outcome prediction of COVID-19 pneumonia. Med Image Anal. 2021;67: 101860.

4. Wynants L, Van Calster B, Collins GS, Riley RD, Heinze G, Schuit E, et al. Prediction models for diagnosis and prognosis of covid-19: systematic review and critical appraisal. BMJ. 2020;369. doi:10.1136/bmj.m1328

5. Messner CB, Demichev V, Wendisch D, Michalick L, White M, Freiwald A, et al. Ultra-High-Throughput Clinical Proteomics Reveals Classifiers of COVID-19 Infection. Cell Syst. 2020;11: 11–24.e4.

6. Shen B, Yi X, Sun Y, Bi X, Du J, Zhang C, et al. Proteomic and Metabolomic Characterization of COVID-19 Patient Sera. Cell. 2020;182: 59–72.e15.

7. Laing AG, Lorenc A, Del Molino Del Barrio I, Das A, Fish M, Monin L, et al. A dynamic COVID-19 immune signature includes associations with poor prognosis. Nat Med. 2020;26: 1623–1635.

8. Liu Y, Gao W, Guo W, Guo Y, Shi M, Dong G, et al. Prominent coagulation disorder is closely related to inflammatory response and could be as a prognostic indicator for ICU patients with COVID-19. J Thromb Thrombolysis. 2020. doi:10.1007/s11239-020-02174-9

9. D’Alessandro A, Thomas T, Dzieciatkowska M, Hill RC, Francis RO, Hudson KE, et al. Serum Proteomics in COVID-19 Patients: Altered Coagulation and Complement Status as a Function of IL-6 Level. J Proteome Res. 2020. doi:10.1021/acs.jproteome.0c00365

10. Schulte-Schrepping J, Reusch N, Paclik D, Baßler K, Schlickeiser S, Zhang B, et al. Severe COVID-19 Is Marked by a Dysregulated Myeloid Cell Compartment. Cell. 2020;182: 1419–1440.e23.

11. Large-Scale Multi-omic Analysis of COVID-19 Severity. Cell Systems. 2021;12: 23–40.e7.

12. Plasma Proteomics Identify Biomarkers and Pathogenesis of COVID-19. Immunity. 2020;53: 1108–1122.e5.

13. Demichev V, Tober-Lau-P Lemke O, Nazarenko T, Thibeault C, Whitwell H, et al. A time-resolved proteomic and prognostic map of COVID-19. accepted Cell Systems. 2021; 1–15.

14. Knaus WA, Draper EA, Wagner DP, Zimmerman JE. APACHE II: a severity of disease classification system. Crit Care Med. 1985;13: 818–829.

15. Ferreira FL. Serial Evaluation of the SOFA Score to Predict Outcome in Critically Ill Patients. JAMA. 2001. p. 1754. doi:10.1001/jama.286.14.1754

16. Wang Z-H, Shu C, Ran X, Xie C-H, Zhang L. Critically Ill Patients with Coronavirus Disease 2019 in a Designated ICU: Clinical Features and Predictors for Mortality. RMHP. 2020;13: 833–845.

17. Zou X, Li S, Fang M, Hu M, Bian Y, Ling J, et al. Acute Physiology and Chronic Health Evaluation II Score as a Predictor of Hospital Mortality in Patients of Coronavirus Disease 2019. Crit Care Med. 2020;48: e657–e665.

18. Thibeault C, Mühlemann B, Helbig ET, Mittermaier M, Lingscheid T, Tober-Lau P, et al. Clinical and virological characteristics of hospitalised COVID-19 patients in a German tertiary care centre during the first wave of the SARS-CoV-2 pandemic: a prospective observational study. Infection. 2021. doi:10.1007/s15010-021-01594-w

19. Wang J, Li D, Dangott LJ, Wu G. Proteomics and Its Role in Nutrition Research. J Nutr. 2006;136: 1759–1762.

20. Elhadad MA, Jonasson C, Huth C, Wilson R, Gieger C, Matias P, et al. Deciphering the Plasma Proteome of Type 2 Diabetes. Diabetes. 2020;69: 2766–2778.

21. Hoogeveen RM, Pereira JPB, Nurmohamed NS, Zampoleri V, Bom MJ, Baragetti A, et al. Improved cardiovascular risk prediction using targeted plasma proteomics in primary prevention. Eur Heart J. 2020;41: 3998–4007.

22. Suhre K, McCarthy MI, Schwenk JM. Genetics meets proteomics: perspectives for large population-based studies. Nat Rev Genet. 2020;22: 19–37.

23. Struwe W, Emmott E, Bailey M, Sharon M, Sinz A, Corrales FJ, et al. The COVID-19 MS Coalition—accelerating diagnostics, prognostics, and treatment. Lancet. 2020;395: 1761–1762.

24. Geyer PE, Kulak NA, Pichler G, Holdt LM, Teupser D, Mann M. Plasma Proteome Profiling to Assess Human Health and Disease. Cell Syst. 2016;2: 185–195.

25. Bruderer R, Muntel J, Müller S, Bernhardt OM, Gandhi T, Cominetti O, et al. Analysis of 1508 Plasma Samples by Capillary-Flow Data-Independent Acquisition Profiles Proteomics of Weight Loss and Maintenance. Mol Cell Proteomics. 2019;18: 1242–1254.

26. Ignjatovic V, Geyer PE, Palaniappan KK, Chaaban JE, Omenn GS, Baker MS, et al. Mass Spectrometry-Based Plasma Proteomics: Considerations from Sample Collection to Achieving Translational Data. J Proteome Res. 2019;18: 4085–4097.

27. Messner CB, Demichev V, Bloomfield N, Yu JSL, White M, Kreidl M, et al. Ultra-fast proteomics with Scanning SWATH. Nat Biotechnol. 2021. doi:10.1038/s41587-021-00860-4

28. Kurth F, Roennefarth M, Thibeault C, Corman VM, Müller-Redetzky H, Mittermaier M, et al. Studying the pathophysiology of coronavirus disease 2019: a protocol for the Berlin prospective COVID-19 patient cohort (Pa-COVID-19). Infection. 2020;48: 619–626.

29. Quan H, Li B, Couris CM, Fushimi K, Graham P, Hider P, et al. Updating and validating the Charlson comorbidity index and score for risk adjustment in hospital discharge abstracts using data from 6 countries. Am J Epidemiol. 2011;173: 676–682.

30. Varol Y, Hakoglu B, Kadri Cirak A, Polat G, Komurcuoglu B, Akkol B, et al. The impact of charlson comorbidity index on mortality from SARS-CoV-2 virus infection and A novel COVID-19 mortality index: CoLACD. Int J Clin Pract. 2020; e13858.

31. Whitwell HJ, Blyuss O, Menon U, Timms JF, Zaikin A. Parenclitic networks for predicting ovarian cancer. Oncotarget. 2018;9: 22717–22726.

32. Krivonosov M, Nazarenko T, Bacalini MG, Franceschi C, Zaikin A, Ivanchenko M. DNA methylation changes with age as a complex system: a parenclitic network approach to a family-based cohort of patients with Down Syndrome. Cold Spring Harbor Laboratory. 2020. p. 2020.03.10.986505. doi:10.1101/2020.03.10.986505

33. Danwang C, Endomba FT, Nkeck JR, Wouna DLA, Robert A, Noubiap JJ. A meta-analysis of potential biomarkers associated with severity of coronavirus disease 2019 (COVID-19). Biomarker Research. 2020;8: 1–13.

34. Henry BM, de Oliveira MHS, Benoit S, Plebani M, Lippi G. Hematologic, biochemical and immune biomarker abnormalities associated with severe illness and mortality in coronavirus disease 2019 (COVID-19): a meta-analysis. Clin Chem Lab Med. 2020;58: 1021–1028.

35. Clinical Characteristics, Treatment, and Outcomes of Critically Ill Patients With COVID-19: A Scoping Review. Mayo Clin Proc. 2021;96: 183–202.

36. Gupta S, Hayek SS, Wang W, Chan L, Mathews KS, Melamed ML, et al. Factors Associated With Death in Critically Ill Patients With Coronavirus Disease 2019 in the US. JAMA Intern Med. 2020;180: 1436–1446.

37. Risitano AM, Mastellos DC, Huber-Lang M, Yancopoulou D, Garlanda C, Ciceri F, et al. Complement as a target in COVID-19? Nat Rev Immunol. 2020;20: 343–344.

38. The hypercoagulable state in COVID-19: Incidence, pathophysiology, and management. Thromb Res. 2020;194: 101–115.

39. Kashuba E, Bailey J, Allsup D, Cawkwell L. The kinin–kallikrein system: physiological roles, pathophysiology and its relationship to cancer biomarkers. Biomarkers. 2013;18: 279–296.

40. van de Veerdonk FL, Netea MG, van Deuren M, van der Meer JWM, de Mast Q, Brüggemann RJ, et al. Kallikrein-kinin blockade in patients with COVID-19 to prevent acute respiratory distress syndrome. 2020 [cited 7 Feb 2021]. doi:10.7554/eLife.57555

41. Colarusso C, Terlizzi M, Pinto A, Sorrentino R. A lesson from a saboteur: High-MW kininogen impact in coronavirus-induced disease 2019. Br J Pharmacol. 2020;177: 4866–4872.

42. van de Veerdonk FL, Kouijzer IJE, de Nooijer AH, van der Hoeven HG, Maas C, Netea MG, et al. Outcomes Associated With Use of a Kinin B2 Receptor Antagonist Among Patients With COVID-19. JAMA Netw Open. 2020;3: e2017708–e2017708.

43. Chao J, Guo Y, Chao L. Protective Role of Endogenous Kallistatin in Vascular Injury and Senescence by Inhibiting Oxidative Stress and Inflammation. Oxid Med Cell Longev. 2018;2018. doi:10.1155/2018/4138560

44. Lin W-C, Chen C-W, Huang Y-W, Chao L, Chao J, Lin Y-S, et al. Kallistatin protects against sepsis-related acute lung injury via inhibiting inflammation and apoptosis. Sci Rep. 2015;5: 12463.

45. NIH ACTIV Trial of blood thinners pauses enrollment of critically ill COVID-19 patients. 22 Dec 2020 [cited 7 Feb 2021]. Available: https://www.nih.gov/news-events/news-releases/nih-activ-trial-blood-thinners-pauses-enrollment-critically-ill-covid-19-patients

46. The REMAP-CAP, Zarychanski R, ACTIV-4a, ATTACC Investigators. Therapeutic anticoagulation in critically ill patients with Covid-19 – preliminary report. bioRxiv. medRxiv; 2021. doi:10.1101/2021.03.10.21252749

47. Zhu X, Wang Y, Zhang H, Liu X, Chen T, Yang R, et al. Genetic Variation of the Human α-2-Heremans-Schmid Glycoprotein (AHSG) Gene Associated with the Risk of SARS-CoV Infection. PLoS One. 2011;6: e23730.

48. Ombrellino M, Wang H, Yang H, Zhang M, Vishnubhakat J, Frazier A, et al. FETUIN, A NEGATIVE ACUTE PHASE PROTEIN, ATTENUATES TNF SYNTHESIS AND THE INNATE INFLAMMATORY RESPONSE TO CARRAGEENAN. Shock. 2001;15: 181.

49. Chua RL, Lukassen S, Trump S, Hennig BP, Wendisch D, Pott F, et al. COVID-19 severity correlates with airway epithelium–immune cell interactions identified by single-cell analysis. Nat Biotechnol. 2020;38: 970–979.

50. SARS-CoV-2 spike protein S1 subunit induces pro-inflammatory responses via toll-like receptor 4 signaling in murine and human macrophages. Heliyon. 2021;7: e06187.

51. Merad M, Martin JC. Pathological inflammation in patients with COVID-19: a key role for monocytes and macrophages. Nat Rev Immunol. 2020;20: 355–362.

52. Team RC, Others. R: A language and environment for statistical computing. Vienna, Austria; 2013. Available: http://cran.univ-paris1.fr/web/packages/dplR/vignettes/intro-dplR.pdf

53. Benjamini Y, Hochberg Y. Controlling the false discovery rate: a practical and powerful approach to multiple testing. J R Stat Soc. 1995. Available: https://rss.onlinelibrary.wiley.com/doi/abs/10.1111/j.2517-6161.1995.tb02031.x

54. Bhowmick P, Mohammed Y, Borchers CH. MRMAssayDB: an integrated resource for validated targeted proteomics assays. Bioinformatics. 2018;34: 3566–3571.

55. Krivonosov M, Nazarenko T, Bacalini MG, Franceschi C. Age-dependent DNA methylation Parenclitic Networks in family-based cohort patients with Down Syndrome. bioRxiv. 2020. Available: https://www.biorxiv.org/content/10.1101/2020.03.10.986505v1.abstract

56. Friedman J, Hastie T, Tibshirani R. Regularization paths for generalized linear models via coordinate descent. J Stat Softw. 2010;33: 1.

