## Supplementary data and machine learning code for "A proteomic survival predictor for COVID-19 patients in intensive care": Parenclitic.R

```
library(stringr)
library(caret)
library(igraph)
library(dplyr)
library(pROC)
library(glmnet)
library(stats)
library(e1071)

Parenclitic <- function(data, features, mode, splittings) {
  rownames(data) <- data$Id
  
  if(mode == "CV") {
    res <- lapply(splittings, function(x) Run_CV(x, data, features))
    
    NETWORKS <- lapply(res, function(x) x[[1]]) 
    CHARACTERISTICS <- lapply(res, function(x) x[[2]]) 
    
    NETWORKS <- do.call("rbind", NETWORKS)
    CHARACTERISTICS <- do.call("rbind", CHARACTERISTICS)
  }
  
  if(mode == "TRAIN_TEST") {
    # edges_cv() is eventually called and performs training only on samples with data$label == 'TRAIN' 
    res <- generate_network(data, features,'MAIN') 
    
    NETWORKS <- res[[1]]
    CHARACTERISTICS <- res[[2]]
  }
  
  return(list(NETWORKS,CHARACTERISTICS))            
}

Run_CV <- function(spl, data, features) {
  data$label = ifelse(data[,spl] == 1, "TRAIN", "TEST")
  generate_network(data, features, spl)
}

generate_network <- function(data, features, RUN_ID) {
  st <- Sys.time()
  print(str_c(RUN_ID,"..."))
  
  list_of_pair <- get_list_of_pair(features)
  edges_dataframe <- lapply(list_of_pair, function(x) 
    edges_cv(x[1], x[2], data))
  
  edges_dataframe <- do.call(rbind, edges_dataframe)
  edges_dataframe <- data.frame(edges_dataframe)
  
  colnames(edges_dataframe) <- c("p1","p2",data$Id)
  for (i in 3:(ncol(edges_dataframe))) edges_dataframe[, i] <- as.numeric(as.character(edges_dataframe[, i]))
  
  NETWORK <- edges_dataframe
  NETWORK$RUN <- RUN_ID

  CHARACTERISTICS <- obtain_characteristics(data, NETWORK[,c("p1","p2",data$Id)], features)
  CHARACTERISTICS$RUN <- RUN_ID
  
  print(Sys.time() - st)
  return(list(NETWORK, CHARACTERISTICS))
}

get_list_of_pair <- function(proteins) {
  list_of_pair <- list()
  k=0
  for(i in 1:(length(proteins)-1)) {
    for(j in (i+1):length(proteins)) {
      k = k + 1
      list_of_pair[[k]] <- c(proteins[i], proteins[j])
    }
  }
  return(list_of_pair)
}

train_svm <- function(data) {
  set.seed(1)
  data$score <- as.factor(data$score)
  result <- svm(score ~ ., data = data, probability = T)
  return(result)
}

prediction <- function(model, data) {
  return(as.numeric(attr(predict(model, data, probability = T), 'probabilities')[,2]))
}

edges_cv <- function(p1,p2,data) {
  df <- data[c(p1, p2, "score", "label")]
  rownames(df) <- data$Id
  df_train <- subset(df,df$label == "TRAIN")[1:(ncol(df)-1)]
  svmfit <- train_svm(df_train)
  return(c(p1, p2, prediction(svmfit, df)))
}

get_graph_object <- function(id, network, proteins) {
  df <- data.frame(p1=network$p1, p2=network$p2,id=network[,id])
  relations <- data.frame(from=as.vector(df$p1), to=as.vector(df$p2))
  g <- graph.data.frame(relations, directed=F, vertices=proteins)
  E(g)$width = ifelse(df$id <= 0, 0.01, df$id)
  return(g)
}

get_graph_object_binary <- function(id, network, proteins) {
  tt <- network[,c("p1", "p2",id)]
  colnames(tt) <- c("p1", "p2","id")
  df <- subset(tt,tt$id > 0.5)
  relations <- data.frame(from=as.vector(df$p1), to=as.vector(df$p2))
  g <- graph.data.frame(relations, directed=F)
  E(g)$width = 1
  return(g)
}

get_characteristics<- function(gr) {
  Eweights <- E(gr)$width
  Eweights_char <- main_stat_char(Eweights)
  names(Eweights_char) <- str_c("Eweights_", names(Eweights_char))
  return(c(Eweights_char))
}

get_characteristics_binary <- function(gr) {
  Counts <- c(length(V(gr)), length(E(gr)))
  names(Counts) <- c('binary_nodes','binary_edges')
  return(c(Counts))
}

obtain_characteristics <- function(data, edges_dataframe, proteins) {
  graph_objects <- lapply(data$Id, function(x) get_graph_object(x,edges_dataframe,proteins))
  graph_objects_binary <- lapply(data$Id, function(x) get_graph_object_binary(x,edges_dataframe,proteins))
  res_characteristics <- data.frame()
  for(j in 1:length(graph_objects)) {
    gr <- graph_objects[[j]]
    gr_b <- graph_objects_binary[[j]]
    res_characteristics <- rbind(res_characteristics, data.frame(t(c(get_characteristics(gr),get_characteristics_binary(gr_b)))))
  }

  res_characteristics$score <- data$score
  res_characteristics$id <- data$Id
  res_characteristics$label <- data$label
  return(res_characteristics)
}

main_stat_char <- function(x) {
  v <- c(max(x), mean(x), sd(x))
  names(v) <- c("max", "mean", "sd")
  return(v)
}

model_on_characteristics <- function(data) {
  set.seed(1)
  return(glmnet(x = as.matrix(data[,-ncol(data)]), y = data$score, family = 'binomial', lambda = 0.01, alpha = 1))
}

char_predict <- function(m, data) {
  return(predict(m, as.matrix(data),type="response"))
}

auc_results <- function(file, loocv_ch, main_ch) {
  runs <- unique(loocv_ch$RUN)
  loocv_results <- c()
  
  features <- colnames(main_ch)[1:(ncol(main_ch) - 4)]
  for(i in 1:length(runs)) {
    temp <- subset(loocv_ch,loocv_ch$RUN == runs[i])
    temp_train <- subset(temp,temp$label == "TRAIN")
    temp_test <- subset(temp,temp$label == "TEST")
    m <- model_on_characteristics(temp_train[,c(features,"score")])
    temp_test$predictions <- char_predict(m, temp_test[,features])
    loocv_results <- rbind(loocv_results, temp_test[,c("predictions","score", "id", "RUN")])
  }
  loocv_results$label = "L3OCV"
  
  temp <- main_ch
  temp_train <- subset(temp,temp$label == "TRAIN")
  temp_test <- subset(temp,temp$label == "TEST")
  
  m <- model_on_characteristics(temp_train[,c(features,"score")])
  main_ch$predictions <- char_predict(m, main_ch[,features])
  main_results <- main_ch[,c("predictions","score", "id", "label")]
  
  loocv_results <- aggregate(predictions ~ id + score + label, loocv_results, FUN = mean)
  if (colnames(loocv_results)[1] != 'predictions') loocv_results = cbind(predictions = loocv_results[,4], loocv_results)[,1:4]

  all_results <- rbind(main_results, loocv_results)
  write.table(all_results, file, sep = '\t', quote = F, row.names=F)
}

nodes_importance <- function(main, main_ch, file) {
  graph_matrix <- lapply(main_ch$id, function(x) ifelse(main[,x] > 0.5,1,0))
  graph_matrix <- do.call(rbind, graph_matrix)
  graph_matrix <- data.frame(graph_matrix)
  colnames(graph_matrix) <- rownames(main)
  rownames(graph_matrix) <- main_ch$id
  graph_matrix <- cbind(graph_matrix,main_ch[,c("score", "id", "label")])
  
  graph_matrix_train <- subset(graph_matrix,graph_matrix$label == "TRAIN" & graph_matrix$score==1) 
  main$mean <- apply(graph_matrix_train[,rownames(main)],2,mean)
  
  proteins <- as.vector(unique(c(as.character(main$p1), as.character(main$p2))))
  mean_gr <- get_graph_object("mean", main, proteins)
  V(mean_gr)$size = strength(mean_gr, weights = E(mean_gr)$width)*10
  
  authority_scores <- authority_score(mean_gr, weights = E(mean_gr)$width)$vector
  df <- data.frame(names=names(authority_scores), authority_scores)
  df <- df[order(df$authority_scores, decreasing = F),]
  write.table(df, file, sep = '\t', quote = F, row.names=F)
}
```
