## Supplementary data and machine learning code for "A proteomic survival predictor for COVID-19 patients in intensive care": PlottingNetworks.R

```
setwd(dirname(rstudioapi::getActiveDocumentContext()$path))
library(cowplot)
library(egg)

plot_network <- function(g, type, title_network, sample, color_vertix) {
  if(type == "weighted") {
    V(g)$size = strength(g, weights = E(g)$width)
    V(g)$color <- rep(color_vertix, length(V(g)))
    E(g)$color <- ifelse(E(g)$width < 0.25, "white",
                         ifelse(E(g)$width < 0.5, "gray",
                                ifelse(E(g)$width < 0.75, "orange","red")))
    
    new_weights_size <- ifelse(E(g)$width < 0.25, 0.25,
                               ifelse(E(g)$width < 0.5, 0.5,
                                      ifelse(E(g)$width < 0.75, 4,6)))
    
    png(filename=str_c("./networks/weighted/",sample,".png"),width = 5000, height = 5000,res=300)
    plot.igraph(g,edge.width = new_weights_size,
                vertex.label.font=1,layout = layout_with_dh, margin=0.25,vertex.label.cex=3)
    title(str_c("W: ",title_network), cex.main = 4)
    dev.off()
  }
  
  if(type == "binary") {
    V(g)$size = strength(g) * 0.5
    V(g)$color <- rep(color_vertix, length(V(g)))
    
    png(filename=str_c("./networks/binary/",sample,".png"),width = 5000, height = 5000,res=300)
    plot.igraph(g,edge.width = 5.0 * E(g)$width,
                vertex.label.font=1,layout =layout_with_dh, margin=0.25,vertex.label.cex=2)
    title(str_c("B: ",title_network), cex.main = 4)
    dev.off()
  }
}


#################
# You need PROTEINS, df and results_TT from Run.R


predictions_Innsbruck <- subset(df, df$label == "TEST")
predictions_Innsbruck <- predictions_Innsbruck[order(predictions_Innsbruck$predictions),]
networks_Innsbruck <- results_TT[[1]][,c("p1","p2",predictions_Innsbruck$id)]

for(i in 1:length(predictions_Innsbruck$id)) {
  sample <- predictions_Innsbruck[i,]$id
  prediction <- predictions_Innsbruck[i,]$predictions
  days <- predictions_Innsbruck[i,]$days
  score <- predictions_Innsbruck[i,]$score
  title = str_c(sample, ", prediction = ",round(prediction,4)," (", score,")")
  
  col_v <- ifelse(i <= 19, "#C0FF3E80", "#FFB90F80")
  g_weighted <- get_graph_object(sample, networks_Innsbruck, PROTEINS)
  plot_network(g_weighted, "weighted", title,sample,col_v)
  
  g_binary = get_graph_object_binary(sample, networks_Innsbruck, PROTEINS)
  plot_network(g_binary, "binary", title,sample,col_v)
}

######### collect weighted plots
plist_weighted <- list()
for(i in 1:length(predictions_Innsbruck$id)) {
  p <- ggdraw() + draw_image(str_c("./networks/weighted/",predictions_Innsbruck[i,]$id,".png"), scale = 1)
  plist_weighted[[i]] <- p
}

######### collect binary plots
plist_binary <- list()
for(i in 1:length(predictions_Innsbruck$id)) {
  p <- ggdraw() + draw_image(str_c("./networks/binary/",predictions_Innsbruck[i,]$id,".png"), scale = 1)
  plist_binary[[i]] <- p
}

######### plotting to pdfs

p_weighted <- ggarrange(plots = plist_weighted, nrow=6,ncol=4)
p_binary <- ggarrange(plots = plist_binary, nrow=6,ncol=4)

ggsave(file="weighted_networks.pdf", p_weighted,width = 20,height=30, units="cm")
ggsave(file="binary_networks.pdf", p_binary,width = 20,height=30, units="cm")
```
