## Supplementary data and machine learning code for "A proteomic survival predictor for COVID-19 patients in intensive care": Prepare data.R

```
setwd(dirname(rstudioapi::getActiveDocumentContext()$path))
library(data.table)
library(stringr)
library(matrixStats)
set.seed(1)

ca.held <- 1 # cases held out each round
ct.held <- 2 # controls held out each round
repeats <- 50 # number of random data partitions to average

gg <- as.data.frame(fread('Charite - log2 protein levels at first max WHO timepoint.tsv',header=T))
rownames(gg) <- gg$Genes
gg$Genes <- NULL
mt <- as.data.frame(fread('Charite - metadata.tsv'))

"
select patients at WHO = 7; exclude patients still critical on 16 Sep 2020; exclude 26 as the death was covid-independent, following recovery to WHO = 4
"
mt <- mt[which(mt$WHO == 7 & mt$I.First.Max.WHO == 1 & mt$Dropout == 0 & (!is.na(mt$Days.Until.Outcome) | mt$Id %in% c(135)) & !(mt$Id %in% c(26))),]

mt <- mt[!duplicated(mt$Id),]
gg <- gg[,!is.na(match(colnames(gg),mt$Id))]
gg <- gg[,match(mt$Id,colnames(gg))]
gg <- as.matrix(gg)

mk <- read.table('Innsbruck - protein levels.tsv',header=T,sep='\t',check.names=F)
rownames(mk) <- mk[,1]
mk[,1] <- NULL
M <- read.table('Innsbruck - metadata.tsv',header=T,sep='\t',check.names=F)
M <- M[match(colnames(mk),M$File.Name),]
M.sel <- which(M$WHO >= 6 & !is.na(M$Died)) # select patients at WHO >= 6
mk <- mk[,M.sel]
M <- M[M.sel,]
mk <- as.matrix(log2(mk))

gg <- gg[!is.na(match(rownames(gg),rownames(mk))),]
mk <- mk[rownames(gg),]

# minimal-value imputation
for (i in 1:nrow(gg)) {
  x <- gg[i,]
  mn <- min(x,na.rm=T)
  if (!is.finite(mn)) mn <- NA
  x[is.na(x)] <- mn
  gg[i,] <- x
  
  x <- mk[i,]
  mn <- min(x,na.rm=T)
  if (!is.finite(mn)) mn <- NA
  x[is.na(x)] <- mn
  mk[i,] <- x
}
gg <- na.omit(gg)
gg <- gg[which(rowSds(gg) > 0),]
mk <- na.omit(mk)
mk <- mk[which(rowSds(mk) > 0),]
gg <- gg[!is.na(match(rownames(gg),rownames(mk))),]
mk <- mk[rownames(gg),]
gg <- na.omit(gg[rownames(mk),])
mk <- na.omit(mk[rownames(gg),])

# standardise the data
means <- rowMeans(gg)
sds <- rowSds(gg)
gg <- (gg - means) / sds
mk <- (mk - means) / sds

mt <- cbind(mt, t(gg))
mt$Id <- paste0('Charite-',mt$Id)
mt$score <- mt$Died
write.table(mt,'Charite samples.tsv',sep='\t',row.names=F,quote=F)

M <- cbind(M, t(mk))
M$Id <- paste0('Innsbruck-',M$Id)
M$score <- M$Died
write.table(M,'Innsbruck samples.tsv',sep='\t',row.names=F,quote=F)

# generate random partitionings for testing the predictor performance
indexCases <- which(mt$Died==1)
indexControls <- which(mt$Died==0)
nca <- length(indexCases)
nct <- length(indexControls)

cases <- sample(indexCases, nca)
controls <- sample(indexControls, nct)
for (i in 1:((nca + nct) * repeats)) {
  cases <- c(cases, sample(indexCases, nca))
  controls <- c(controls, sample(indexControls, nct))
}

res <- list()
lct <- 0
lca <- 0
while(lca < nca * repeats | lct < nct * repeats) {
  res[[length(res)+1]] <- c(cases[(lca + 1) : (lca + ca.held)], controls[(lct + 1) : (lct + ct.held)])
  lca <- lca + ca.held
  lct <- lct + ct.held
}

samples <- data.frame(id=mt$Id, score=mt$Died)
for(i in 1:length(res)) {
  v <- rep(1, nrow(mt))
  df <- data.frame(samp = v, id=mt$Id)
  positions <- as.character(res[[i]])
  df[positions,]$samp  <- rep(0, length(positions))
  colnames(df) <- c(str_c("RUN",i), "id")
  samples = merge(samples, df, by="id")
}
write.table(samples,'Splittings.tsv',sep='\t',row.names=F,quote=F)
```
