## Supplementary data and machine learning code for "A proteomic survival predictor for COVID-19 patients in intensive care": Run.R

```
setwd(dirname(rstudioapi::getActiveDocumentContext()$path))
library(data.table)
library(stringr)
library(matrixStats)
set.seed(1)

#source("Prepare data.R") # uncomment to generate the input data
source("Prepare data.R")
source("Parenclitic.R")

# Proteins which (i) have MRM assays available (ii) have >=3 peptides detected
PROTEINS <- c("A2M", "AGT", "AHSG", "ALB", "AMBP", "APOA1", "APOB", 
              "APOE", "APOH", "B2M", "BTD", "C1QA", "C1QB", "C1QC", 
              "C3", "C4A", "C4B", "C5", "CP", "CRP", "CST3", "F10", 
              "F11", "F12", "F13A1", "F13B", "F2", "F5", "F9", "FGA", 
              "FGB", "FN1", "HBA1", "HBB", "HP", "HPX", "KLKB1", "LPA", 
              "LYZ", "ORM1", "ORM2", "PF4", "PLG", "PPBP", "PROS1", 
              "RBP4", "S100A8", "S100A9", "SERPINA1", "SERPINC1", 
              "SERPINF2", "SERPING1", "SHBG", "TF","TFRC", "TTR", "VWF")

splittings <- read.table('Splittings.tsv', stringsAsFactors=F, sep = "\t", header = T)
splittings_column_names <- setdiff(colnames(splittings),c("X","id","score"))
rownames(splittings) <- splittings$id

DATA_TRAIN <- as.data.frame(fread('Charite samples.tsv', stringsAsFactors=F, sep = "\t", header=T))  
rownames(DATA_TRAIN) <- DATA_TRAIN$Id
DATA_FOR_L3OCV <- cbind(DATA_TRAIN, splittings[rownames(DATA_TRAIN),splittings_column_names])

DATA_TEST <- as.data.frame(fread('Innsbruck samples.tsv', stringsAsFactors=F, sep = "\t", header=T))  
rownames(DATA_TEST) <- DATA_TEST$Id
DATA_TRAIN$label = "TRAIN"
DATA_TEST$label = "TEST"
common <- intersect(colnames(DATA_TRAIN), colnames(DATA_TEST))
PROTEINS <- PROTEINS[PROTEINS %in% common]
print('Proteins used to build the model:')
print(PROTEINS)

# Form the data for training on Charite/testing on Innsbruck
DATA_FOR_TRAIN_TEST <- rbind(DATA_TRAIN[,common],DATA_TEST[,common])

results_L3OCV <- Parenclitic(DATA_FOR_L3OCV, PROTEINS, "CV", splittings_column_names)
results_TT <- Parenclitic(DATA_FOR_TRAIN_TEST, PROTEINS, "TRAIN_TEST", c())

auc_results('results.tsv', results_L3OCV[[2]], results_TT[[2]])
nodes_importance(results_TT[[1]], results_TT[[2]], 'protein importance.txt')

# Plots
set.seed(4) # for jitter
df <- as.data.frame(fread('results.tsv'))
df$days = 0
df$days <- DATA_TRAIN$Days.Until.Outcome[match(df$id,DATA_TRAIN$Id)]
df$days[is.na(df$days)] <- DATA_TEST$Days.Until.Outcome[match(df$id[is.na(df$days)],DATA_TEST$Id)]
cv <- df[(df$label == 'L3OCV'),]
test <- df[(df$label == 'TEST'),]

# For survival analysis
library(survival)
library(broom)
margin <- 0.678
cv$group <- as.integer((cv$predictions > margin))
cv$days[is.na(cv$days) | cv$score == 0] <- max(cv$days[cv$score == 1],na.rm=T)
f <- Surv(cv$days, cv$score) ~ cv$group
fit.cv <- survfit(f)
p.cv <- signif(broom::glance(survdiff(f,rho=0))$p.value, 2)

test$group <- as.integer((test$predictions > margin))
test$days[is.na(test$days) | test$score == 0] <- max(test$days[test$score == 1],na.rm=T)
f <- Surv(test$days, test$score) ~ test$group
fit.test <- survfit(f)
p.test <- signif(broom::glance(survdiff(f,rho=0))$p.value, 2)

pdf('plots.pdf')
par(mfrow=c(3,3), font.main = 1)
rc <- roc(cv$score, cv$predictions, direction = '<')
cx <- 1.5
cx.lgd <- 1.1
plot(rc, main = paste0('Charite cohort, AUC = ',signif(as.numeric(rc$auc),2)),cex.main = cx,cex.lab=cx,cex.axis=cx)

p <- wilcox.test(cv$predictions ~ as.factor(cv$score))$p.value[1]
fc <- as.factor(unlist(list('0'='no','1'='yes')[as.character(cv$score)]))
boxplot(cv$predictions ~ fc, main = paste0('Charite cohort\nP = ',signif(p,2)),cex.main = cx,cex.lab=cx,cex.axis=cx,range=0, col = c('grey','darkorange'),xlab = 'Died', ylab = 'Predicted probability',ylim=range(0,1))
points(cv$predictions ~ jitter(cv$score+1,0.5), pch = 16, col = "#000000A0", cex = 1.0)

plot(fit.cv, col = c('black','darkorange'), xlab = 'Days', ylab = 'Survival', main = paste0('P = ',p.cv), cex.main=cx,cex.lab=cx,cex.axis=cx,lwd = 1.5)
legend(-5, 0.35, legend=c(paste0("Prediction < ",margin), paste0("Prediction > ",margin)), col=c("black", "darkorange"), cex=cx.lgd, box.lty=0, lwd = 1.5)

rc <- roc(test$score, test$predictions, direction = '<')
plot(rc, main = paste0('Innsbruck cohort, AUC = ',signif(as.numeric(rc$auc),2)),cex.main = cx,cex.lab=cx,cex.axis=cx)

p <- wilcox.test(test$predictions ~ as.factor(test$score))$p.value[1]
fc <- as.factor(unlist(list('0'='no','1'='yes')[as.character(test$score)]))
boxplot(test$predictions ~ fc, main = paste0('Innsbruck cohort\nP = ',signif(p,2)),cex.main = cx,cex.lab=cx,cex.axis=cx,range=0, col = c('grey','darkorange'),xlab = 'Died', ylab = 'Predicted probability',ylim=range(0,1))
points(test$predictions ~ jitter(test$score+1,0.5), pch = 16, col = "#000000A0", cex = 1.0)

plot(fit.test, col = c('black','darkorange'), xlab = 'Days', ylab = 'Survival', main = paste0('P = ',p.test), cex.main=cx,cex.lab=cx,cex.axis=cx,lwd = 1.5)
legend(-4, 0.35, legend=c(paste0("Prediction < ",margin), paste0("Prediction > ",margin)), col=c("black", "darkorange"), cex=cx.lgd, box.lty=0, lwd = 1.5)

# APACHE, SOFA and Charlson
df <- as.data.frame(fread('Charlson_APACHE_SOFA.txt'))

df$x <- df$APACHE
p <- wilcox.test(df$x ~ as.factor(df$I.Died))$p.value[1]
fc <- as.factor(unlist(list('0'='no','1'='yes')[as.character(df$I.Died)]))
boxplot(df$x ~ fc, main = paste0('P = ',signif(p,2)),cex.main = cx,cex.lab=cx,cex.axis=cx,range=0, col = c('grey','darkorange'),xlab = 'Died', ylab = 'APACHE II score')
points(df$x ~ jitter(df$I.Died+1,0.5), pch = 16, col = "#000000A0", cex = 1.0)

df$x <- df$I.SOFA
p <- wilcox.test(df$x ~ as.factor(df$I.Died))$p.value[1]
fc <- as.factor(unlist(list('0'='no','1'='yes')[as.character(df$I.Died)]))
boxplot(df$x ~ fc, main = paste0('P = ',signif(p,2)),cex.main = cx,cex.lab=cx,cex.axis=cx,range=0, col = c('grey','darkorange'),xlab = 'Died', ylab = 'SOFA score')
points(df$x ~ jitter(df$I.Died+1,0.5), pch = 16, col = "#000000A0", cex = 1.0)

df$x <- df$I.Charlson
p <- wilcox.test(df$x ~ as.factor(df$I.Died))$p.value[1]
fc <- as.factor(unlist(list('0'='no','1'='yes')[as.character(df$I.Died)]))
boxplot(df$x ~ fc, main = paste0('P = ',signif(p,2)),cex.main = cx,cex.lab=cx,cex.axis=cx,range=0, col = c('grey','darkorange'),xlab = 'Died', ylab = 'Charlson index')
points(df$x ~ jitter(df$I.Died+1,0.5), pch = 16, col = "#000000A0", cex = 1.0)

dev.off()

df <- as.data.frame(fread('results.tsv'))
source("PlottingNetworks.R")
```
